# Performance and health economic evaluation of the Mount Sinai COVID-19 serological assay identifies modification of thresholding as necessary to maximise specificity of the assay

**DOI:** 10.1101/2020.06.11.20128306

**Authors:** Stuart A Rushworth, Benjamin B Johnson, Karen Ashurst, Rose Davidson, Paige Paddy, Jayna J Mistry, Jamie A Moore, Charlotte Hellmich, Dylan R Edwards, Daniela Da Luz Afonso, Georgios Xydopoulos, Richard Goodwin, Javier Gomez, Reenesh Prakash, Samir Dervisevic, Richard Fordham, Kristian M Bowles, James G W Smith

## Abstract

We evaluated the FDA approved SARS-CoV-2 immunoassay (developed at Mount Sinai, by Krammer and colleagues) for the identification of COVID-19 seroconversion and potential cross-reactivity of the assay in a United Kingdom (UK) National Health Service (NHS) hospital setting. In our ‘set up’ cohort we found that the SARS-CoV-2 IgG was detectable in 100% of patients tested 14 days post positive COVID-19 nucleic acid test. Serum samples taken from pregnant women in 2018 were used as a negative control group with zero false positives. We also analysed samples from patients with non-COVID-19 viral infections, paraproteinaemia or autoantibodies and found false positive results in 6/179. Modification of the sensitivity threshold to five standard deviations from the mean of the control group eliminated all false positive result in the ‘set up’ cohort. We confirmed the validity of the test with a revised threshold on an independent prospective ‘validation cohort’ of patient samples. Taking data from both cohorts we report a sensitivity of the Mount Sinai assay of 96.6% (28/29) and specificity of 100% (299/299) using a revised threshold cut-off, at a time point at least 14 days since the diagnostic antigen test. Finally, we conducted a health economic probabilistic sensitivity analysis (PSA) on the costs of producing the tests, and the mean cost we estimate to be 13.63 pounds sterling (95%CI 9.63 - 18.40), allowing its cost effectiveness to be tested against other antibody tests. In summary, we report that the Mount Sinai IgG ELISA assay is highly sensitive test for SARS-Cov-2 infection, however modification of thresholding was required to minimise false positive results.

## Introduction

Coronavirus disease 2019 (COVID-19) is an infectious disease caused by severe acute respiratory syndrome coronavirus 2 (SARS-CoV-2). Common symptoms include fever, cough, and shortness of breath(1). However, reports from China suggest that as many as four fifths of infected people may have been asymptomatic (2). In Iceland and Po (Italy) approximately 40% of infections were reported to be asymptomatic (3, 4). Given that the majority of cases result in a mild or subclinical acute phase, then it is likely the estimations of incidence are an under-estimate. Furthermore, in the UK diagnostic testing has to date largely been limited to mainly symptomatic individuals and primarily those admitted to hospitals. Taken together, the vast majority of people who have had COVID-19 infection in the UK are highly likely to have been unconfirmed by laboratory antigen testing.

At present, it is not known if or to what degree previous COVID-19 infection confers protection on virus re-exposure. However, if COVID-19 infection generates immunity to subsequent COVID-19 infection, or abrogates the severity of the illness on re-exposure, then evidence of prior infection may have important healthcare and social implications for individuals in the future. To address the need to identify previous infection, a number of serological assays have been developed (5). Serological analysis of patients following COVID-19 infection demonstrates that circa 99% of people develop a B cell response (6, 7). In addition, the FDA approved (8) ELISA assay developed at Mount Sinai by Krammer and colleagues appears to correlate well with a neutralization assay (9). Therefore, taken together these data suggest that the great majority of infected people subsequently make neutralizing antibodies, even if the infection was mild. The clinical implications of these observations remain at present largely undefined.

It has long been recognised that immunoassays may be subject to positive and negative interference by both polyclonal and monoclonal antibodies, which can then impact assay performance (10, 11). Report of a viral antibody detection ELISA assay, assessed at the time of the 2003 SARS-corona virus (SARS-CoV) outbreak (12), described false-positive SARS-CoV results occurring as a consequence of autoantibodies in 19/58 patients with systemic lupus erythematosus (SLE) (13). Similarly, paraproteins (which occur in myeloma and lymphoma, and commonly in older people as a monoclonal gammopathy of undetermined significance (MGUS), have been reported to interfere in immunoassays (14). In addition, antibody testing will need to be able to differentiate between SARS-CoV-2 and other members of the Coronaviruses group as well as other non-COVID-19 viral infections. In general, both sensitivity and specificity should be as high as possible (5). Accordingly, as antibody testing is expected to play an important and widespread role in managing the COVID-19 pandemic, then assessment and understanding of the impacts of non-COVID-19 viral infections, and polyclonal and monoclonal antibodies on assay performance are required.

We evaluated and validated the Mount Sinai ELISA assay (9, 15), on sets of serum samples from UK patients following SARS-CoV-2 antigen confirmed COVID-19 infection and compared these data to the assay results of serum from patients with confirmed non-COVID19 viral infection, a bank of historic stored serum samples from 2018 (predating the emergence of COVID-19) and samples from patients with paraproteinaemia and autoantibodies. Finally, we conducted a health economic probabilistic sensitivity analysis (PSA) to estimate the mean cost of producing the test result.

## Methods

Expression vectors for the SARS-CoV-2 receptor-binding domain (RBD) and SARS-CoV-2 full-length spike protein were kindly provided by the Krammer laboratory (Icahn School of Medicine at Mount Sinai, New York, USA) and, ELISA plates were produced and assayed according to the detailed published protocol (9). An overview of these methods is provided below.

### ELISA plates

Plasmid DNA was transformed into chemically competent E.coli (ThermoFisher Scientific), and isolated using an endotoxin-free maxiprep kit (Macherey Nagel). Plasmid DNA was transfected into HEK293F’ cells (ThermoFisher Scientific) and Histagged protein were isolated 72 hours post transfection through gravity flow purification. Isolated protein was quantified using a Pierce BCA assay (ThermoFisher Scientific), diluted to a concentration of 2 µg/ml in PBS, and used to coat 4 HBX ELISA plates (ThermoFisher Scientific) overnight at 4°C. Reactivity of plates to SARS-CoV antibody was confirmed through the use of the monoclonal antibody CR30229 (16) (Absolute Antibody) (Supplementary figure 1).

### Human serum samples

This study was sponsored by the Norfolk and Norwich University Hospital NHS Trust (UK) and conducted under ethical approval of the University of East Anglia Faculty of Medicine and Health Research Ethics Committee (UK). Left over and stored material from blood samples, taken in routine clinical practice, at the Norfolk and Norwich University Hospital, and which was no longer required for clinical use, was used in these assays.

### ELISA assay

ELISA plates were washed 3 times with PBS containing 0.1% tween 20 (PBS-T), followed by a 1 hour blocking incubation in PBS-T containing 1% milk powder (w/v). Serum samples were first added to RBD coated plates at a final sample dilution of 1:50 in PBS-T containing 1% milk powder (w/v). Following a 2 hour incubation at room temperature, the ELISA plates were washed three times with PBS-T and incubated for 1 hour (± 5 minutes) with HRP-labelled anti-human Fab specific secondary IgG, IgA, or IgM antibody (Sigma), diluted 1:3000 in PBS-T containing 1% milk powder (w/v). After the incubation, the ELISA plates were washed again three times with PBS-T and colour development achieved by incubating with SIGMA*FAST*^*™*^ OPD solution (Sigma) (prepared following manufacturer’s instructions). The colour development reaction was stopped by addition of 3M hydrochloric acid to all wells, exactly 10 minutes after the addition of the o-phenylenediamine dihydrochloride (OPD) solution (Sigma), and absorbance of 490 nm was recorded using a FluoStar Omega plate reader (BMG Labtech). Samples that exceeded OD490 cut-off of three standard deviations (SD) of the negative control group were assigned presumptive positive. The presumptive positive samples were taken to the second step of the assay protocol where they underwentthe same procedure but at a reciprocal dilution against full-length spike protein coated plates. Samples that exceeded OD490 cut-off of three standard deviations of the negative control group at two consecutive dilutions were assigned positive.

### Validation in an NHS laboratory

The Norfolk and Norwich University Hospital (NNUH) provides acute NHS Trust tertiary level specialist care and laboratory services to a catchment area of approximately 1 million people in the East of England. The laboratory holds United Kingdom Accreditation Service (UKAS) laboratory accreditation (17) and the NNUH Virology department is compliant with ISO15189.

The ELISA protocol was scripted on the DYNEX DS2 Automated ELISA System (Dynex Technologies) and used to screen previously untested sera samples from negative control patients with non-COVID19 viral infection, paraproteinemia and autoantibodies, and test patients reported as PCR positive for COVID19 at a time point at least 14 days since the diagnostic confirmatory antigen test.

### Health Economic Analysis

In order to calculate the unit cost of this antibody test, each step was broken down into a how much time it is necessary to conduct it and what are the necessary staff, equipment and consumables. The unit cost of blood sample, GP appointment and A&E visits were taken from the NHS Reference Costs and the Personal Social Service Research Unit (18). The costs for lab technicians were not available in these reports and were collected individually from pay scale websites (19). The hourly costs for staff were modified depending on the duration of the task. The estimation of the cost of using the relevant equipment was performed by calculating the annual depreciation of the equipment factored by the percentage of time that is required to process the samples for the new test. Apart from the deterministic outcomes, a probabilistic sensitivity analysis was implemented to reduce the variance in the dataset and define the confidence intervals.

## Results

### Evaluation of the Mount Sinai ELISA assay on sera from confirmed COVID-19 PCR positive samples and negative control serum taken in 2018 – ‘Set up cohort’

A group of 50 presumed healthy negative samples (pregnant women pre-2019), were screened against a group of 50 samples previously confirmed positive for SARS-CoV-2 antibodies from patients who had previously tested positive for SARS-CoV-2 using the AusDiagnostics PCR test (20). ELISAs were initially performed at a sera dilution of 1:50 for IgG, IgA and IgM against SARS-CoV-2 receptor-binding domain coated plates. Results show a significant increase level of SARS-CoV-2 IgG, IgA and IgM antibody in the PCR positive COVID-19 group compared to the control negative serum that had been taken from pregnant women in 2018 (Figure 1A and Supplementary Figure 2; (P<0.0001)). Positive threshold was defined, as previously described by the Mount Sinai team, using the suggested threshold of 3 standard deviations from the mean of the negative control group. Using this threshold, analysis of the serum from patients previously diagnosed with COVID-19 infection identified SARS-CoV-2 spike protein antibodies in 41/50 samples for IgG, 38/50 samples for IgA and 31/50 samples for IgM (Figure 1A). These 41 samples are considered ‘presumptive positive’ following this Receptor Binding Domain of SARS-CoV-2 (RBD) step of the Mount Sinai assay at this stage, and were then taken forward to a second confirmatory step of the assay protocol. On the second stage reciprocal dilution, against full-length spike protein coated plates, confirmed 39/50 samples to be SARS-CoV-2 IgG positive (Figure 1B).

**Figure 1.**
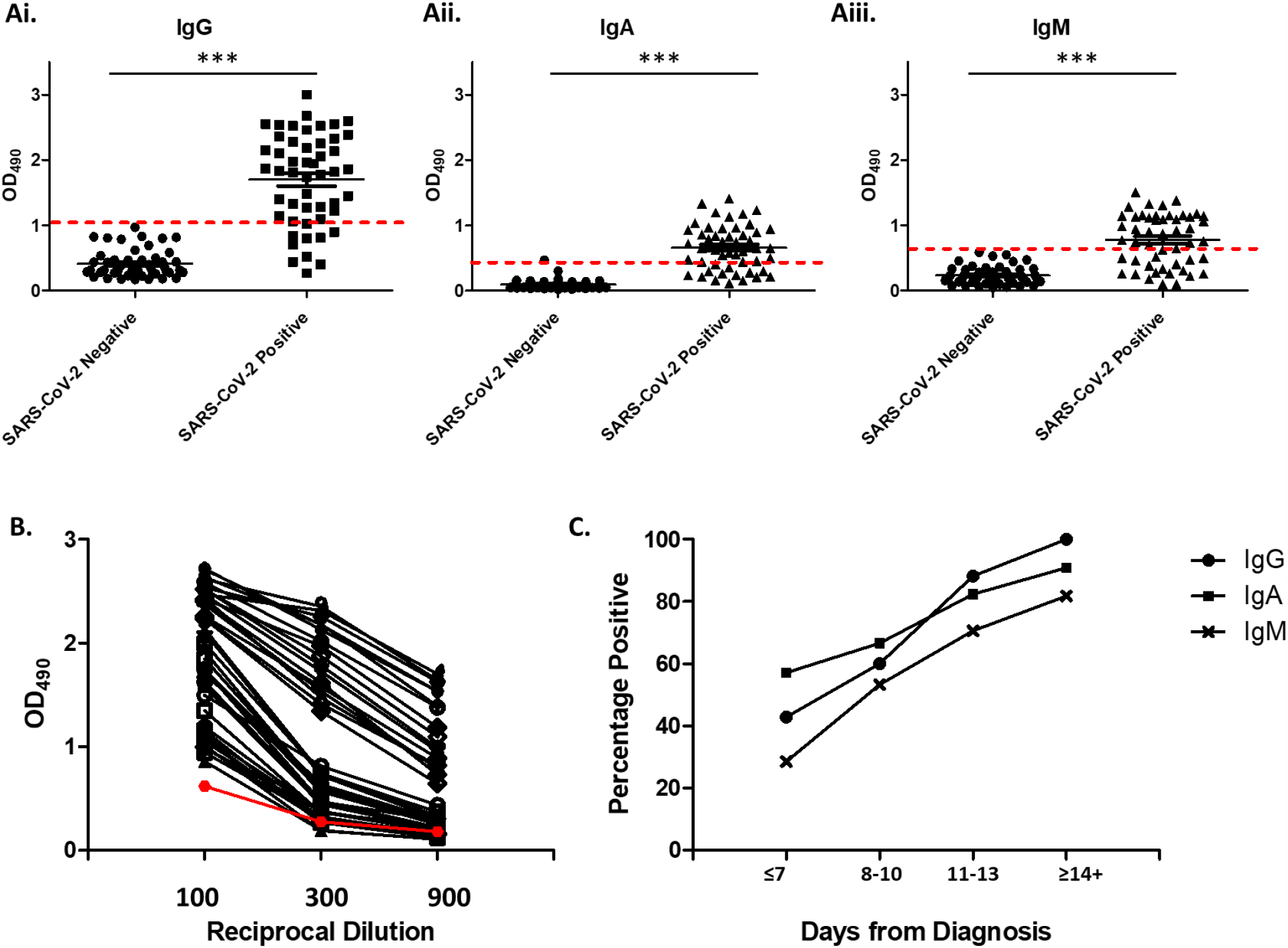
Reactivity of control and SARS-CoV-2 convalescent sera to spike antigens. **A)** Primary screen of 50 SARS-CoV-2 negative samples (pregnant women pre-2019)and 50 SARS-CoV-2 positive samples (COVID19 positive AusDiagnostics PCR test), against RBD of spike protein at a single 1:50 dilution using IgG, IgA and IgM secondary antibodies (threshold of 3 standard deviations of negative controls shown as dotted red line). Statistical analyses were performed using an unpaired two-tailed Student’s t-test in GraphPad Prism. P<0.0001. **B)** Presumed positives from IgG primary screen serially diluted against full length spike protein (threshold of 3 standard deviations of negative controls shown as red line). **C)** The percentage of positive samples within SARS-CoV-2 positive group, plotted against the day from PCR diagnosis.

Emergence of a detectable antibody response is time dependent, and because the natural history of the infection is known to be variable and that the onset of symptoms was thought to be a subjective timepoint, we next plotted antibody results against the time since diagnostic PCR positive test against COVID19 seroconversion result (days). Figure 1C shows all sera samples taken greater than 14 days post PCR test were positive for IgG (11/11), but this was not the case for IgA (10/11) or IgM (9/11) SARS-CoV-2 antibody. Together these data confirm the Mount Sinai assay to be highly sensitive for seroconversion of IgG when the test is performed over 14 days following the diagnostic PCR test.

### Evaluation of cross-reactivity in the Mount Sinai ELISA assay– ‘Set up cohort’

It is recognised that other infectious illness, as well as polyclonal and monoclonal antibodies, can impact ELISA assay performance. Therefore, we investigated whether the Mount Sinai SARS-CoV-2 IgG assay would have cross-reactivity in patients with other medical conditions. Firstly, sera from patients with non-COVID-19 viral infections (including seasonal coronavirus and influenza) was tested. We selected sera stored in our NHS regional diagnostic virology laboratory taken between 1^st^ January 2019 and 27th November 2019. As, Covid-19 emerged in Wuhan, China in December 2019 (21), the first UK case was diagnosed week commencing 27th January 2020 in Yorkshire (22), and the first case of Covid-19 in our own area was diagnosed on 6^th^ March 2020, we considered these samples to be from patients who had not been exposed to SARS-CoV-2. Figure 2A shows that 42/47 samples from this group were established as negative for SARS-CoV-2 IgG antibody in the first RBD screening test step, and 5/47 required confirmatory assessment with the second dilution assay. We also ran the same assay using secondary antibodies to human IgA and IgM. The IgA test was negative in 44/47 and IgM negative in 44/47 after step 1. Following the reciprocal dilution step, 4/5 of the ‘presumed positive’ samples assayed were confirmed as IgG ‘positive’ by this test (Supplementary Figure 3A).

**Figure 2.**
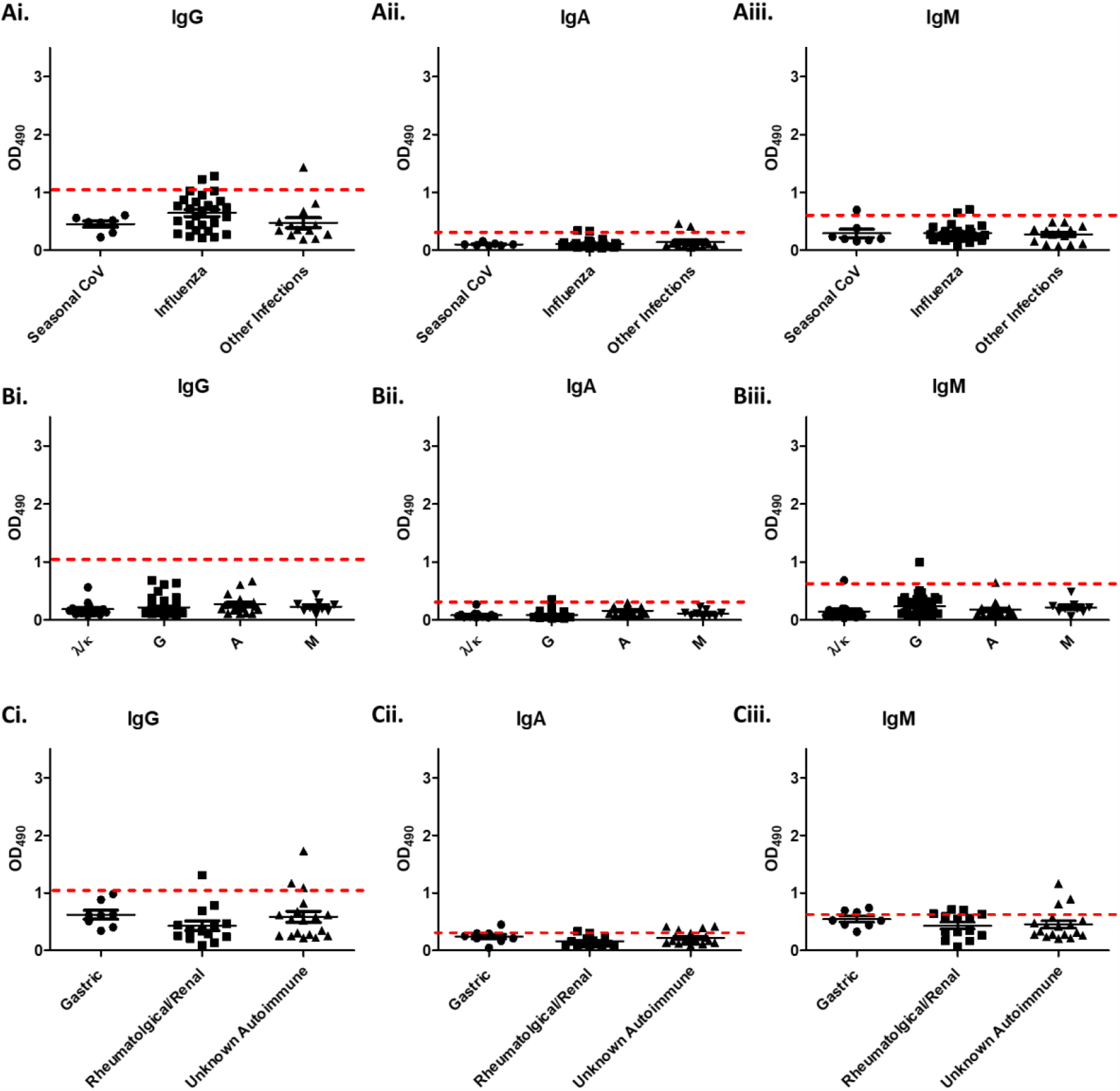
Reactivity interference by viral infections, paraproteins, and autoantibodies. Primary screen of sera from patients with **A)** viral infections, including seasonal corona viruses, influenza and other viral infections, **B)** paraproteinaemia, including kappa and lambda light chain paraproteins (λK), IgG paraproteins (G), IgA paraproteins (A), and IgM paraproteins, and **C)** autoantibodies, including gastric autoimmune diseases, rheumatalogical/renal autoimmune diseases, and unknown autoimmune disease.

Next, we screened sera from 92 patients with paraproteinaemia and 40 patients with autoantibodies for potential ‘cross-reactivity’ in the Mount Sinai ELISA. These blood samples were taken from patients in May 2020. As the blood tests were taken for other medical reasons and the background incidence of COVID-19 in our region at the time was thought to be low, we expected a low rate of SARS-COV-2 positivity in this group. None of the 92 samples from patients with paraproteinaemia were presumed positives for IgG, IgA or IgM (Figure 2B) after the RBD step of the assay and were confirmed as negative for SARS-CoV-2 IgG antibody. There were however ‘presumed positives’ following the RBD step in the autoantibody samples group: 4/40 for IgG, 5/40 for IgA, and 8/40 for IgM (Figure 2C). These were taken forward to the reciprocal dilution test, after which 2/4 IgG ‘presumed positives’ were confirmed as positive by this assay (Supplementary Figure 3B). Although these blood test were taken to look for auto-antibody, not Covid-19 infection, these samples were taken during a period of time when Covid-19 was known to be present in our community. These data suggest the IgG Mount Sinai assay may have some potential cross-reactivity with sera from some patients with autoimmune diseases.

Following the identification of p*otential* false positives within the cross-reactive groups, we sought to determine if the specificity of the Mount Sinai assay could be increased with a more stringent threshold. The threshold was therefore increased from 3 SD to 5 SD of the mean of the negative control. On re-analysis at this new threshold only 1 of the 9 presumed positives samples from the original primary RBD screen (where 3 SD was used as a cut off) remained positive at a level above the cut-off (Figure 3A). Furthermore, this sample was not subsequently above the 5 SD threshold for two consecutive dilutions in the secondary reciprocal dilution test (Figure 3B). Therefore, by moving the cut-off to 5 SD from the mean of the normal controls there were no false positive results in the samples from patients with paraproteinaemia, non-COVID-19 viral infection and autoimmune disease groups.

**Figure 3.**
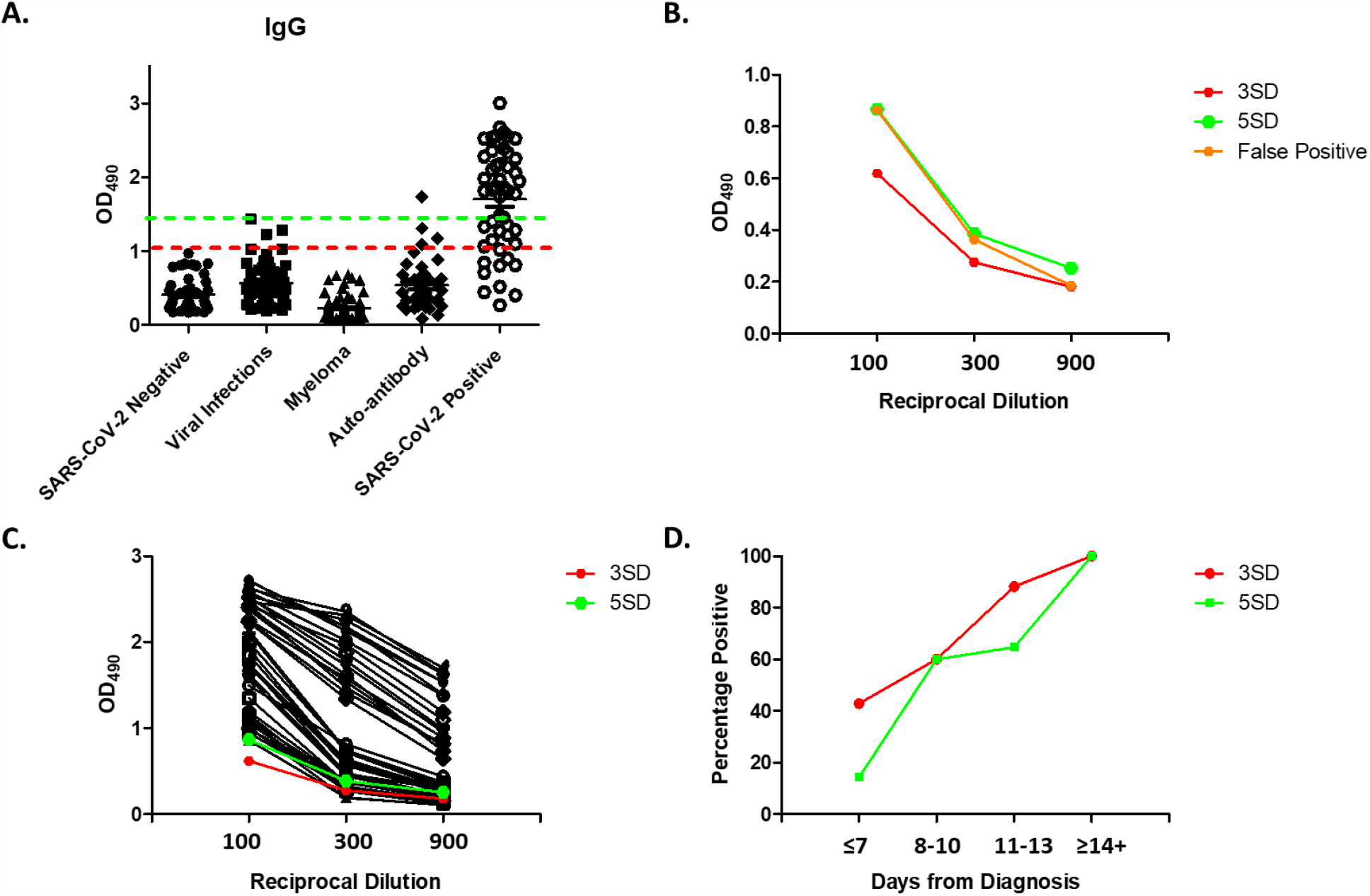
Increased threshold from 3 to 5 standard deviations of the negative controls. **A)** Primary screen of 50 SARS-CoV-2 negative (pregnant women pre-2019), 50 SARS-CoV-2 positive (COVID19 positive AusDiagnostics PCR test), 47 viral infection, 92 myeloma, and 40 autoantibody sera samples against RBD of spike protein at a single 1:50 dilution. Original threshold of 3 standard deviations (3SD) shown as red dotted line and increased threshold of 5 standard deviations (5SD) shown as green dotted line. **B)** Remaining false positive serially diluted against full length spike protein. **C)** Presumed positives from SARS-CoV-2 positive group serially diluted against full length spike protein. **D)** The percentage of positive samples within SARS-CoV-2 positive group, plotted against the day from PCR diagnosis for the original 3SD and increased 5SD thresholds.

However, on reanalysis of all of the patients who had previously tested positive by Sars-Cov-2 antigen PCR the increased 5 SD cut-off threshold reduced the number of positive results above the threshold from 39/50 to 32/50 (Figure 3A). These 32 samples all remained above the 5 SD threshold in the secondary reciprocal dilution against the full-length spike protein (Figure 3C). Figure 3D shows the seven samples no longer considered positive as a result in the change of the threshold were all from earlier time points during the course of the illness. Even with the revised 5 SD threshold the assay identified all serum samples taken greater than 14 days post PCR test being positive.

These data show that although we have identified some potential cross-reactivity at the 3 SD threshold, resulting in false positive results in patients with non-COVID-19 viral infection and autoimmune disease, the false positives could be eliminated through the use of an increased assay cut-off threshold. This increased threshold still allowed successful identification of all samples from patients greater than 14 days from the diagnostic PCR test.

### Validation of Mount Sinai assay in a United Kingdom (UK) National Health Service (NHS) hospital setting – ‘Validation cohort’

Having set a revised cut-off threshold in the ‘setup’ cohort of patients, we next prospectively tested the assay in an independent ‘validation cohort’ set of patients. Furthermore, the assay protocol and reagents were provided to an NHS service laboratory to validate its use in an NHS clinical diagnostic environment. A previously untested group of 72 negative control samples from patients with non-COVID19 viral infection, paraproteinaemia and autoantibodies was screened along with previously untested group of 18 samples from patients reported as PCR positive for COVID19 at a time point at least 14 days since the diagnostic confirmatory antigen test. The non-Covid-19 viral infection samples were from samples taken before November 27^th^ 2019. All other blood samples were taken in May and June of 2020.

Figure 4A shows the NHS laboratory identified SARS-CoV-2 IgG antibodies in 17/18 samples tested. On testing of the control group, 70/72 patient samples were identified as being negative for SARS-CoV-2 IgG antibody following the RBD step of the assay using the 5 SD threshold. These two samples were in the autoantibody group taken in May 2020. All 19 presumed positives from the primary RBD screen (17 in the known COVID-19 group and 2 in the control group), were confirmed positive in the second stage reciprocal dilution assay (Figure 4B).

**Figure 4.**
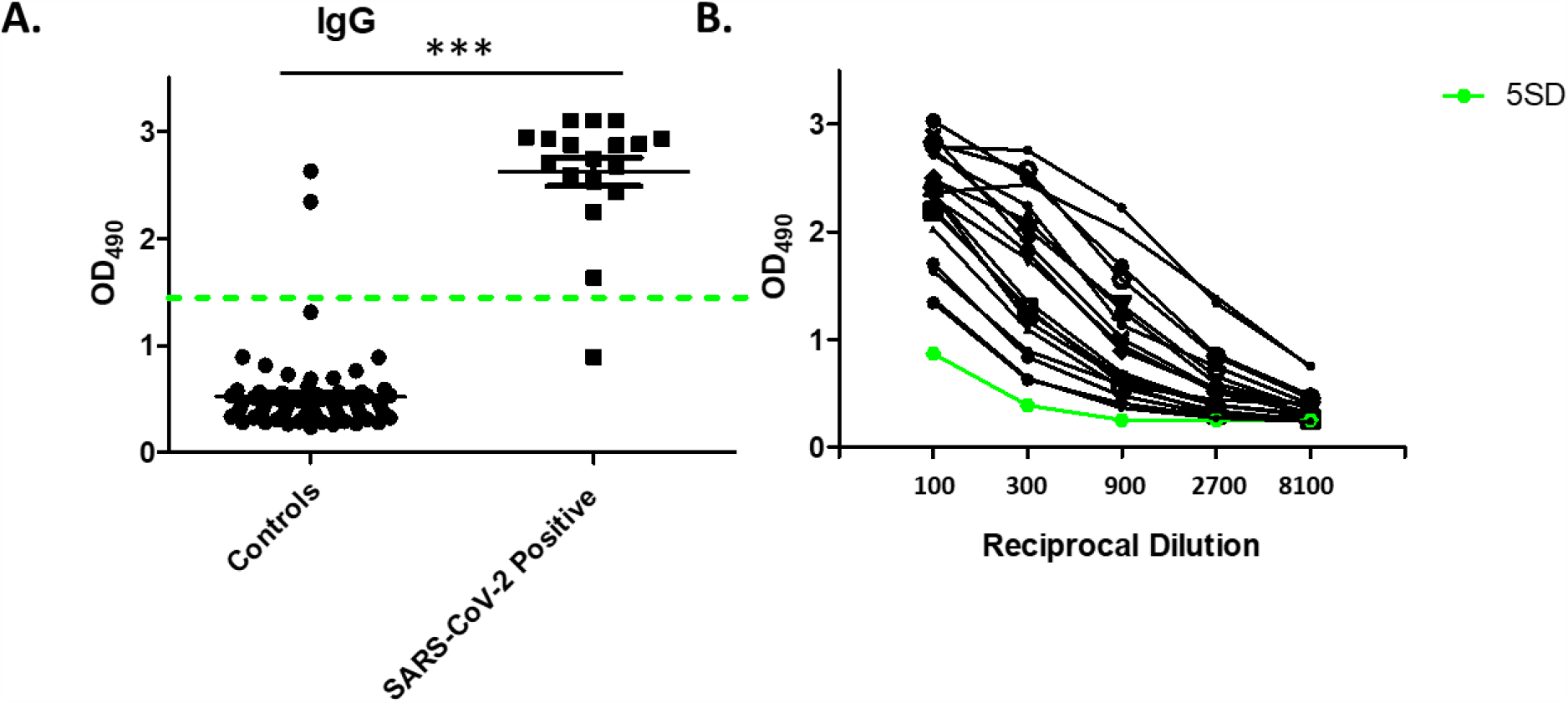
Validation of Mount Sinai assay in a United Kingdom (UK) National Health Service (NHS) hospital setting. **A)** Primary screen of a previously untested group of 72 negative control samples from patients with non-COVID19 viral infection, paraproteinaemia and autoantibodies, and a previously untested group of 18 samples from patients reported as PCR positive for COVID19. Threshold of 5 standard deviations (5SD) shown as green dotted line. **B)** 17 Presumed positives from SARS-CoV-2 positive group and 2 from control group serially diluted against full length spike protein. Threshold of 5 standard deviations (5SD) shown as green line.

To further understand the results from the two unexpected Mount Sinai assay SARS-CoV-2 IgG antibody positive patient samples from our auto-antibody control group, we next performed the SARS-CoV-2 IgG antibody assay using the same serum on our Abbott SARS-CoV-2 IgG system. We found that the serum samples from both patients which had tested positive in the Mount Sinai assay also tested positive for SARS-CoV-2 IgG antibody on the Abbott antibody assay as well (data not shown). The Abbott assay is a chemiluminescent microparticle assay manufactured by Abbott Laboratories and is listed as CE marked and approved for antibody testing in the UK (23). We now consider both these patients to have had previously undiagnosed Covid-19 infection and this 2/72 (3%) positive SARS-COV-2 antibody rate is consistent with our estimated community prevalence of Covid-19 infection at the time the blood samples were taken. Together, these data show that the Mount Sinai assay is highly sensitive for detection of SARS-CoV-2 IgG antibody in an NHS setting.

### Health Economic Analysis

In order to establish a reliable price per test a probabilistic sensitivity analysis (PSA) was undertaken on the cost of materials and salaries etc. used in producing the ELIZA test. This is done to assure the robustness of estimates under uncertainty, such as the unit prices of raw materials and resources costs and their likely variance. Probabilistic sensitivity analysis (PSA) is a technique used formally in economic modelling that allows the specification of a statistical level of confidence in the outputs of the analysis, in relation to the expected uncertainty in the model inputs. These are represented as distributions around each estimated known value. A Gamma distribution (skewed but always positive) of costs was assumed as is conventional with input variables such as costs which in the case of medical treatments are not normally distributed. A set of input parameters is drawn by random sampling from known distributions, and the model generates outputs (cost and health outcome) which are repeated in many iterations of the model (typically 1000–10,000). This results in a distribution of outputs that can be graphed in a scatterplot presenting the price per test per model iteration, and analysed (Supplementary Figure 4A and B). Uncertainty surrounding the fixed cost data was examined using sensitivity analysis and the mean cost was shown be £13.63 (95%CI £9.63 - £18.40), which is approximately $17 (US dollar) or €15 (Euro) at the time of writing (supplementary document 1).

### Summary

Collectively our findings confirm the high sensitivity of the Mount Sinai SARS-CoV-2 IgG antibody ELISA assay in a UK NHS clinical setting. In our ‘set-up cohort’ we found that moving the positive result cut-off from 3 standard deviations of the mean of the control samples to 5 standard deviations of the mean of the control samples was necessary to optimise assay performance. We confirmed this observation in an independent ‘validation cohort’. In the validation cohort two patients were found to have SARS-CoV-2 IgG antibodies, both of whom had previously been investigated as inpatients for possible COVID-19 and found to have been negative at the time for SARS-CoV-2 antigen by PCR. These two samples were then tested on the UK government approved Abbott ‘ARCHITECT *i* System’ SARS-CoV-2 antibody test (23), and on balance the clinical assumption was made that these patients had previously had undiagnosed COVID-19 infection, so were removed from our final control group. Therefore, taking data from both cohorts we report a sensitivity of the Mount Sinai assay of 96.6% (28/29) and specificity of 100% (299/299) using a revised threshold cut-off, at a time point at least 14 days since the diagnostic antigen test. This data compares favourably with assessments of other RBD and non-RBD based assays (24)

For the costing of this novel testing approach we have used well established NHS costing reports (18) for the primary care and hospital appointments and administration costs. All costs and resource use relate to laboratory consumables were provided by the scientific team and laboratory technician experts. The probabilistic analysis provided the confidence intervals around the price of the newly developed test, hopefully reducing the variance around the costs from different sources and institutions that could be found around the UK. The estimation of the cost of using the relevant equipment was performed by calculating the annual depreciation of the equipment factored by the percentage of time that is required to process the samples for the new test. When considering the potential future scale of antibody testing, here we provide a bench mark ‘per test’ cost for the Mount Sinai assay in a UK NHS setting, against which other similar assays can be compared in a transparent fashion.

Multiple SARS-COV-2 antibody assays have been developed and/or validated in a growing number of different individual labs. Generally these tests show an improved sensitivity with N antigen-based tests, that IgG tests perform better when compared to IgM tests, and most show better sensitivity when the samples are taken longer after the onset of symptoms (25). However, because even the same assay in routine clinical practice will be conducted in different laboratories, it will be essential to calibrate and standardize the assay delivered by different laboratories by using well-defined standard references as part of diagnostic assay validation. Thus, setting up reference panels and quality assurance programs will be essential components in the establishment and delivery of widespread laboratory performance management systems (26).

To conclude, here we report that the Mount Sinai IgG ELISA assay is highly sensitive and apparent cost-effective test for SARS-Cov-2 infection in a UK NHS acute hospital laboratory setting. Individual laboratory optimisation of thresholding determined by local control material is recommended.

## Data Availability

The datasets generated during and/or analysed during the current study are available from the corresponding author on reasonable request.

## Acknowledgements

J.G.W.S. is supported by the Academy of Medical Sciences/the Wellcome Trust/ the Government Department of Business, Energy and Industrial Strategy/the British Heart Foundation/Diabetes UK Springboard Award [SBF005\1057]. S.A.R is funded by The Rosetrees Trust and the Big C charity. Development of SARS-CoV-2 reagents was partially supported by the NIAID Centers of Excellence for Influenza Research and Surveillance (CEIRS) contract HHSN272201400008C.

## Supplementary Figures

**Supplementary Figure 1.**
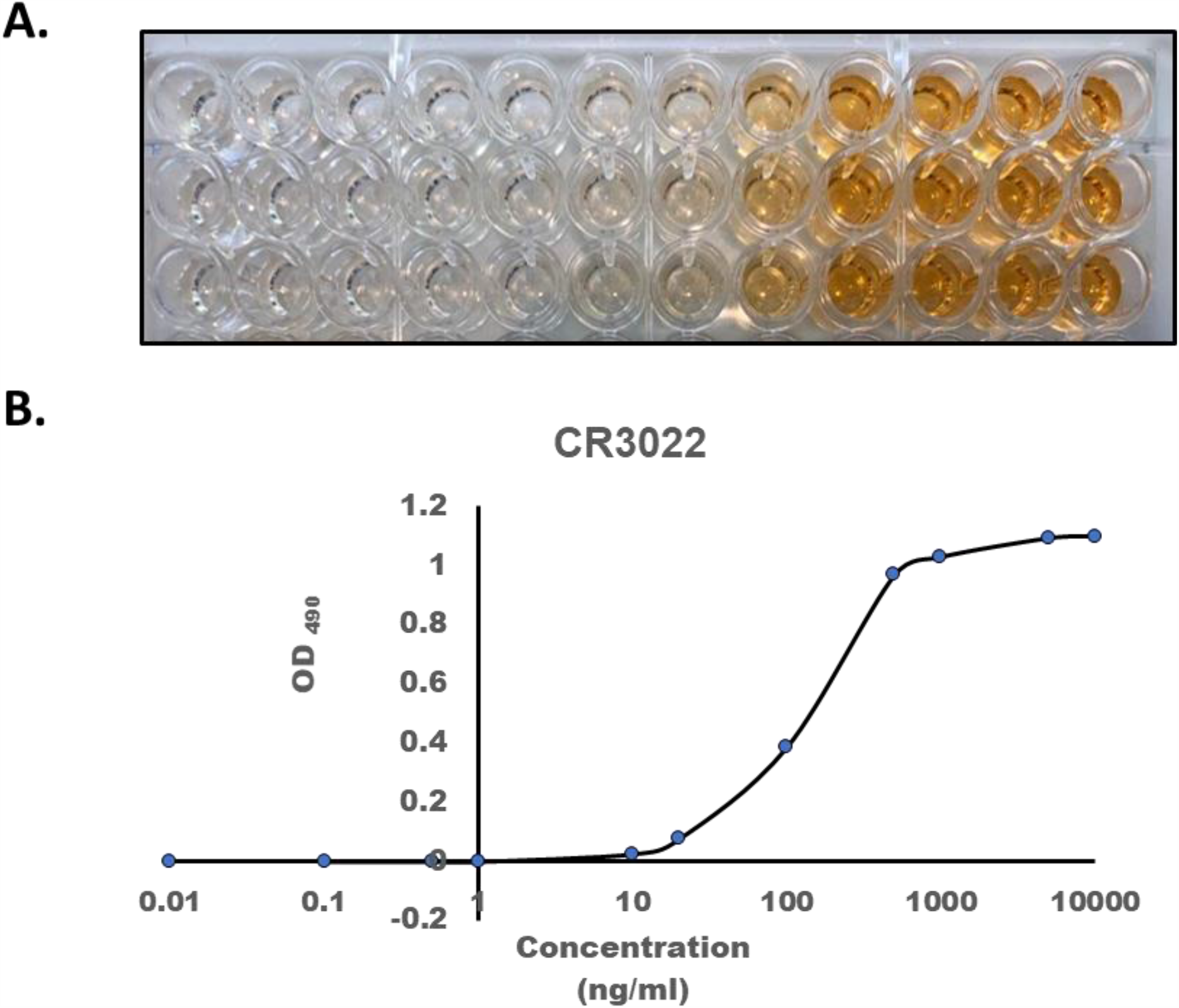
Reactivity of monoclonal CR30229 antibody to spike antigen. **A)** Photograph of RBD coated ELISA plate reacted with CR3022 antibody at a concentration range 0.01 – 10,000 ng/ml, with **B)** Corresponding 490 nm absorbance values.

**Supplementary Figure 2.**
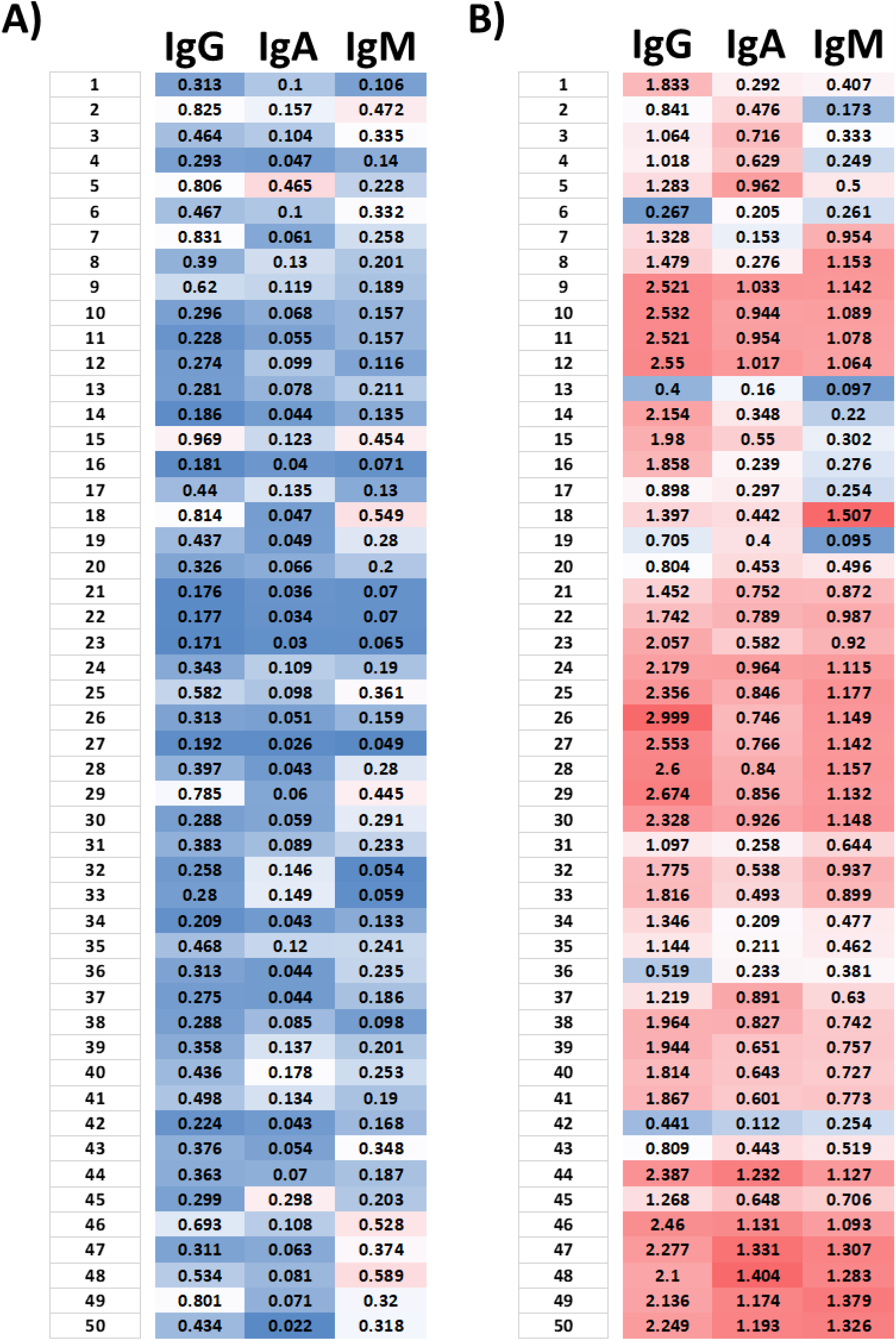
Reactivity of control and SARS-CoV-2 convalescent sera to spike antigens. Raw 490 nm absorbance values of **A)** 50 SARS-CoV-2 negative samples (pregnant women pre-2019), and **B)** 50 SARS-CoV-2 positive samples (COVID-19 positive AusDiagnostics PCR test).

**Supplementary Figure 3.**
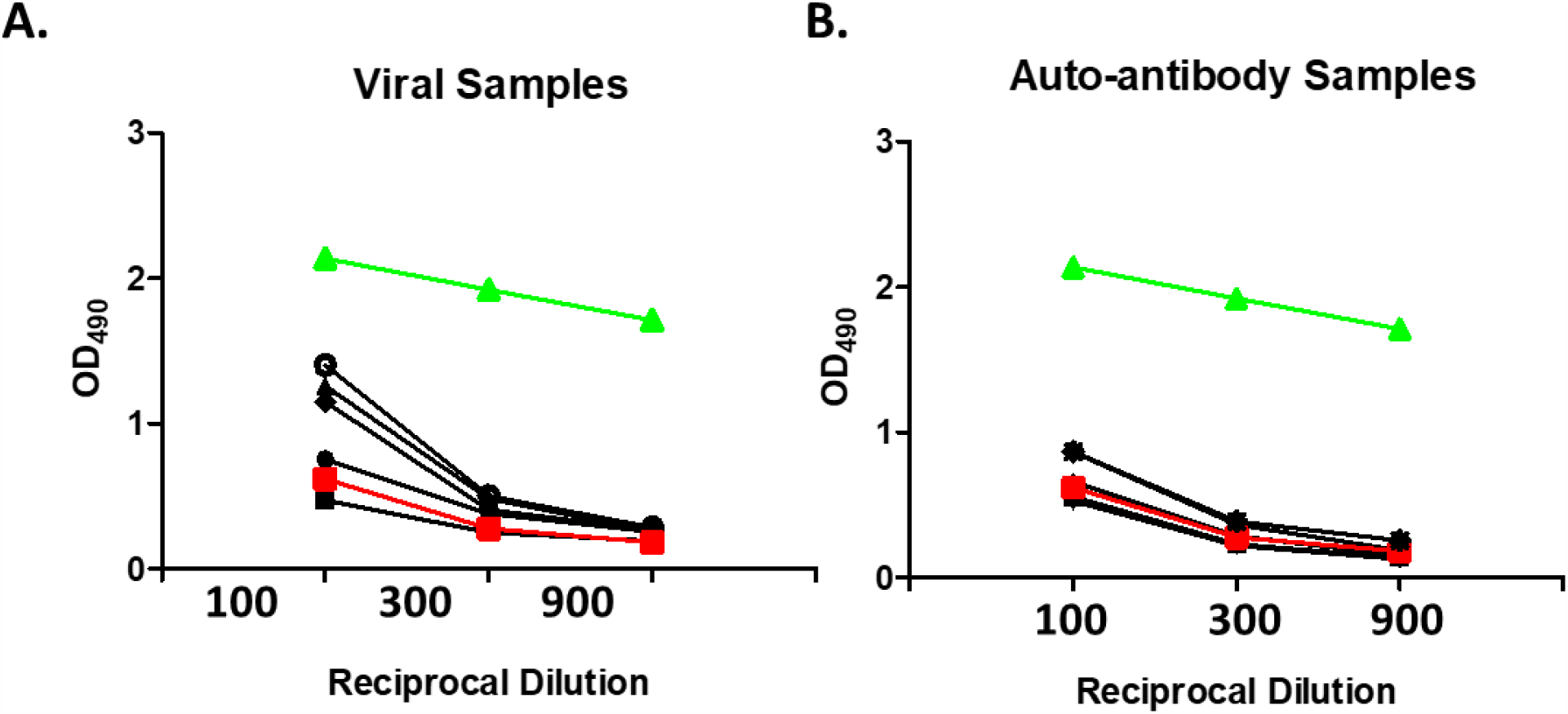
Reactivity interference by viral infections and autoantibodies. Reciprocal dilution of presumed positive sera from patients with **A)** viral infections and **B)** autoimmune disease. Threshold of three standard deviations of the negative control group shown as red line. Positive control sample shown as green line.

**Supplementary Figure 4.**
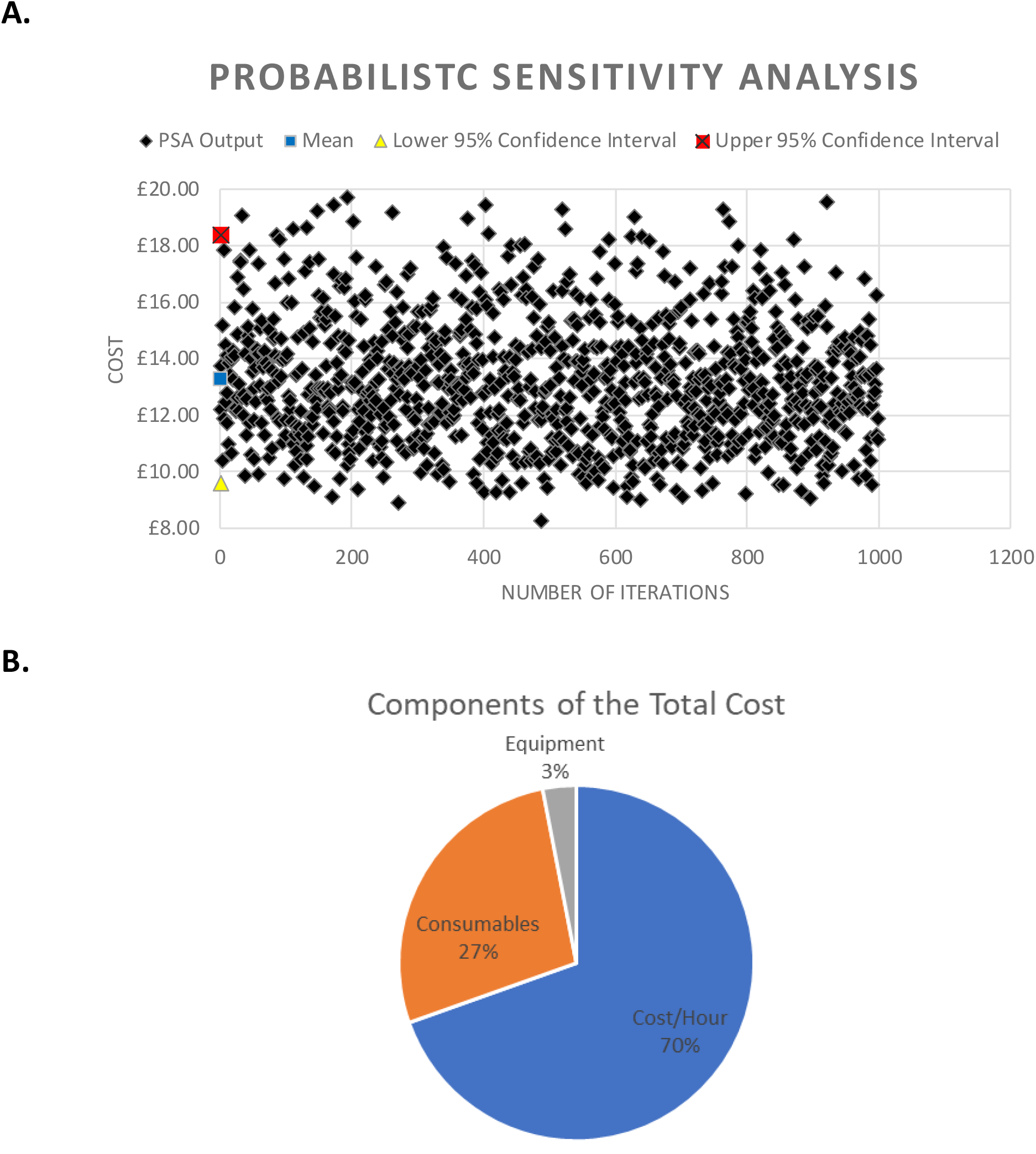
Probabilistic sensitivity analysis estimations of costs. A. Values were denser around the mean (£13.30, 95% Confidence Interval £9.63-£18.40). B. Components of total costs.

